# Protocol for a Cross-Sectional Comparative Analysis of Condom Use and STI Prevalence Among PrEP Users and Non-Users at a Specialised STI Wellness Clinic in Eswatini

**DOI:** 10.64898/2026.05.04.26352404

**Authors:** Yves Mafulu, Victor Williams, Phumlile Ndlovu, Khetsiwe L. Maseko, Sandile Ndabezitha, Nkosana Ndlovu, Siphesihle Gwebu, Nomcebo Matsenjwa, Benedictus Deku, Ncedo Mhlanga, Nkululeko Dube

## Abstract

1.

**Introduction:** Sexually transmitted infections (STIs) represent a significant public health challenge globally, with the Southern Africa region experiencing particularly high rates. In Eswatini, the burden of STIs, including HIV, is alarming, necessitating effective prevention strategies. Pre-exposure prophylaxis (PrEP) has been introduced as a key intervention, yet its impact on condom use and STI prevalence remains underexplored.

**Objective:** This study aims to evaluate and compare condom use patterns and the prevalence of STIs among clients using PrEP and those who do not, among clients attending an STI Wellness Clinic in Eswatini.

**Methods:** A prospective cross-sectional quantitative study will be conducted from February to June 2026 at the LaMvelase STI Wellness Centre. Participants will include HIV-negative individuals aged 15 and above, recruited through purposive sampling. Data will be collected via structured questionnaires and medical records, assessing condom use frequency, sexual behaviour, and STI rates. Laboratory testing will be conducted to confirm STI diagnoses. Statistical analyses will include descriptive statistics and logistic regression to identify associations between PrEP use and STI prevalence.

**Results:** This protocol describes a study designed to assess the relationship between PrEP use, condom use behaviour, and STI prevalence among clients attending a specialised STI clinic in Eswatini. Findings will inform public health strategies and educational programs aimed at reducing STI rates and improving sexual health outcomes in Eswatini.

**Conclusion:** Understanding the dynamics between PrEP use and sexual health practices is crucial for optimising STI prevention strategies. This research will contribute valuable data to guide interventions and health policies and to design more effective intervention strategies in high STI prevalence settings, ultimately supporting efforts to mitigate the impact of STIs and HIV in Eswatini.

## 2. Introduction

### 2.1. Background

Sexually transmitted infections (STIs) remain a significant public health issue worldwide. The WHO estimates that over 1 million STIs, including chlamydia, gonorrhoea, syphilis, and trichomoniasis, among others, are contracted daily (WHO, 2024). Untreated STIs can lead to serious health complications, including infertility, increased risk of HIV transmission, and complications during pregnancy[1,2]. The Southern Africa region has one of the highest rates of STIs, and estimates indicate that approximately 70% of new HIV infections globally occur in sub-Saharan Africa [3]. In response to this crisis, various preventive measures, including the use of condoms and pre-exposure prophylaxis (PrEP), have been implemented to curb the spread of STIs and HIV.

In Eswatini, the burden of STIs, including HIV/AIDS, remains alarmingly high. In a study assessing the acceptability of point-of-care testing (POCT) for Chlamydia trachomatis (CT) and Neisseria gonorrhoea (NG) piloted among sexually active adults aged 18-45 years, 22% were found to be infected[4]. In another study testing for NG, CT and Trichomonas Vaginalis (TV) among adolescents and young people living with HIV in Eswatini, 15.5% of people aged 15-24 years tested positive, and 25% among females aged 20-25 years[5]. Ginidza et al. (2017) found a prevalence of 19.4% among females aged 15-49 years. This is particularly concerning given the country’s high HIV prevalence rate, which is one of the highest in the world. The 2021 Swaziland HIV Incidence Measurement Survey (SHIMS) 3 reveals that the national HIV prevalence among adults aged 15 years and older is approximately 24.8%(Eswatini Ministry of Health, 2023).

PrEP is a biomedical intervention that involves taking Antiretroviral therapy (ART) to prevent the acquisition of HIV [7]. It has been demonstrated that PrEP with ART is effective in reducing the risk of acquiring HIV in people at risk[8]. The United States Food and Drug Administration (FDA) approved the fixed-dose combination tablet of Tenofovir Disoproxil fumarate (TDF)/Emtricitabine (FTC) for use as PrEP for adults in 2012[8]. In Eswatini, the introduction and expansion of PrEP have been integral to the country’s successful HIV response. Since the WHO recommended PrEP in 2015, Eswatini has progressively implemented and scaled up this prevention method since 2016, and Eswatini uses Tenofovir Disoproxil fumarate (TDF)/Lamivudine (3TC) as PrEP. By 2019, oral PrEP was introduced widely across public and private healthcare facilities, with notable increases in both the number of facilities offering PrEP and the number of users(WHO Eswatini, 2023).

The Ministry of Health (MoH) of Eswatini, supported by WHO and various partners, has expanded PrEP options to include event-driven PrEP for men and is currently piloting the Dapavirine Vaginal Ring and injectable PrEP. This expansion has contributed to a significant decline in HIV incidence, moving Eswatini closer to its goal of eliminating AIDS as a public health threat by 2030 (WHO Eswatini, 2023). However, the efficacy of PrEP in preventing HIV transmission does not negate the necessity of other preventive measures, such as condom use, which remains a critical component of STI and HIV prevention strategies(Freeborn and Portillo, 2018).

Different studies in other settings are showing varying impacts of PrEP on the use of condoms. Some studies suggest that PrEP use may result in reduced condom use due to perceived decreased risk, a phenomenon called sexual risk compensation, others found that PrEP use may result in maintaining or even increasing condom use due to an increased awareness of the risk of STI and need for prevention [10–12]. Understanding these dynamics is necessary to optimise STI prevention strategies and improve health outcomes. This manuscript reports the study protocol prior to initiation of participant recruitment and data collection.

### 2.2. Justification for the study

In Eswatini, few studies have been conducted to investigate the dynamics between PrEP and condom use patterns. It is necessary to determine whether the introduction of PrEP affects condom use among individuals using PrEP compared to PrEP non-users. This is crucial for the optimisation of sexual health interventions, especially in settings with high STI rates, like in Eswatini. This study is justified by the need to evaluate how the use of PrEP influences condom use behaviours and STI incidence. Despite PrEP’s effectiveness in preventing HIV(CDC, 2020), its impact on sexual health practices, including condom use, remains largely underexplored. This research intends to fill this knowledge gap by providing insights into the perceived STI risks associated with PrEP use.

### 2.3. Potential use/relevance of study findings

The findings of this study can guide public health strategies, help refine educational programs, and enhance intervention methods tailored to reduce STI prevalence and improve sexual health outcomes. Furthermore, it addresses the unique needs of Eswatini’s population, where STI rates and HIV prevalence are significant, thus contributing to more targeted and effective health policies.

## 3. Aim and Objectives

### 3.1. Study question

1. Is the use of HIV Pre-exposure Prophylaxis (PrEP) associated with an increased prevalence of Sexually Transmitted Infections (STIs)?
2. Is the use of HIV Pre-exposure Prophylaxis (PrEP) associated with decreased condom use among clients attending the STI Wellness Clinic?

### 3.2. Aim

To describe condom use patterns and prevalence of sexually transmitted infections (STIs) among PrEP users and non-users at the LaMvelase STI Wellness Clinic in Eswatini.

### 3.3. Objectives

1. To describe the frequency and consistency of condom use between PrEP users and PrEP-naïve clients.
2. To determine the prevalence of STIs among PrEP users and PrEP-naïve users attending the STI wellness clinic in different population groups or categories.
3. To describe demographic and behavioural factors that may influence condom use and STI rates among PrEP users and non-users in the study population.
4. To determine the incidence and prevalence of HIV in clients attending the STI wellness clinic.

### 3.4. Definition of Key Terms

The following are key concepts and terms defined or explained within the context of the current study:

- **Pre-exposure prophylaxis:** A biomedical intervention that involves taking Antiretroviral therapy (ART) to prevent the acquisition of HIV [7]. In Eswatini, the regimen used is Tenofovir Disoproxil fumarate (TDF)/Lamivudine (3TC), Dapivirine vaginal ring, and Long-Acting Cabotegravir (CAB-LA). The country will soon introduce Lenacapavir, which is new in the market.
- **Sexually Transmitted Infections (STIs):** previously known as sexually transmitted diseases (STDs), involve the transmission of an organism between sexual partners through different routes of sexual contact, either oral, anal, or vaginal. The most common STIs include gonorrhoea, chlamydia, syphilis, trichomonas, herpes viruses, and human papillomavirus (HPV) [13].
- **Risk Compensation**: is a behavioural and psychological theory that refers to the behavioural adjustments individuals make in response to perceived improvements in safety, such as those resulting from regulations or design enhancements [14]. In our study context, the phenomenon suggests that PrEP use may result in reduced condom use due to a perceived decrease in HIV risk[10]. Some studies, however, indicate that PrEP use may result in maintaining or even increasing condom use.
- **Condom Use Consistency:** in the context of the current study, measures the frequency of condom use during sexual intercourse (e.g., always, sometimes, never) and the types of sexual activity where condoms are used.
- **PrEP Users (PrEP-Experienced):** individuals who are currently using or have used HIV pre-exposure prophylaxis (PrEP) as part of their HIV prevention strategy for at least 1 month.
- **Non-PrEP Users (PrEP-Naïve):** individuals who have never used HIV PrEP before enrollment in the study.
- **Point of Care Testing (POCT):** rapid diagnostic tests performed at the site or time of patient care.
- **Event-Driven PrEP**: a PrEP dosing strategy where medication is taken around the time of sexual activity rather than daily (e.g., 2-1-1 method for men).

## 4. Methodology

### 4.1. Study design

This study will use a prospective cross-sectional design to describe condom use patterns and the prevalence of STIs between HIV PrEP users and non-PrEP users in clients accessing services at the LaMvelase STI Wellness clinic from February to June 2026. Data for the study will be obtained from a structured survey of patients and their medical records. Data will include information on condom use, sexual behaviour and STIs.

### 4.2. Study Status and Timeline

At the time of submission of this protocol, the study has not yet commenced, and no data collection or data analysis has been conducted. Participant recruitment is expected to begin in February 2026 and will be completed by June 2026. Data collection will occur concurrently with recruitment and is expected to be completed by June 2026. Data cleaning and analysis are anticipated to take place between June and July 2026. Study results are expected from August 2026 onwards.

### 4.3. Study setting

The study will be conducted at the LaMvelase STI Wellness Centre, which operates within the LaMvelase Centre of Excellence, a standalone facility specialising in HIV care, serving approximately 15,000 registered patients. The STI Wellness Centre focuses on STIs and provides STIs screening, diagnosis, and treatment, and offers prevention commodities such as male and female condoms, lubricants, PrEP, PEP, and referrals for voluntary medical male circumcision (VMMC). Psychosocial support is also available to help clients manage the emotional aspects of sexual health. Open to both HIV-positive and HIV-negative individuals, the Wellness Centre aims to serve sexually active people at higher risk for HIV and other STIs, including sex workers and other key populations.

### 4.4. Study population and participants

The study population will consist of PrEP users and non-PrEP users who present at the Wellness facility for HIV and STI services. All HIV-negative individuals are routinely offered post-test HIV preventive services at the Wellness Clinic and will be potential participants in the study.

### 4.5. Sampling and Sample Size

A systematic random sampling will be used to recruit participants in the PrEP/non-PrEP cohort. The sampling frame will be all eligible clients attending the clinic. A sampling interval (k) will be determined based on client load. Every k-th eligible client will be approached for recruitment until the target sample size is met. Potential participants who meet the inclusion criteria will be informed about the research and offered an opportunity to participate. Information on the study will include the objectives, associated risks of participating, and possible benefits. Those who agree to participate will be required to sign an informed consent form before they can be included.

The sample size will be calculated using the formula for cross-sectional studies:

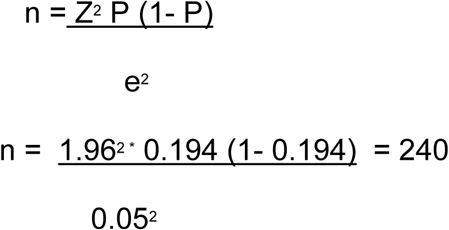

Where:

N = the required sample size

Z = the standard deviation at 95% confidence level (1.96).

P = Estimated prevalence of the outcome or proportion (expressed as a decimal). The researchers could not find current STI prevalence in Eswatini; however, Ginidza et al. (2017) reported that the prevalence of STIs in Eswatini is 19.4% (0.194).

e =the standard error (0.05).

Therefore, the minimum sample size for this study is 240. After accounting for 10% of potential refusal, the anticipated sample size will be 264.

### 4.6. Inclusion and exclusion criteria

#### 4.6.1. Inclusion criteria

In the PrEP/non-PrEP

I. HIV-negative clients aged 15 years and above, regardless of gender, who came for HIV preventive services.
II. Clients must have attended the Wellness Clinic for STI management during the study period.
III. **HIV PrEP Status**: Participants must fall into one of the following categories:

- **HIV PrEP Users (PrEP-experienced)**: Individuals who are currently using or have used HIV pre-exposure prophylaxis (PrEP) as part of their HIV prevention strategy for at least 1 month.
- **Non-PrEP Users (PrEP-naive)**: Individuals who have never used HIV pre-exposure prophylaxis (PrEP) at any time before enrollment in the study.

#### 4.6.2. Exclusion criteria

I. HIV-positive clients, regardless of age and gender. Exclusion of this group of the population is justified by the fact that the study focuses on clients on PrEP who are supposed to be HIV negative.
II. HIV-negative clients less than 15 years of age.
III. Individuals who are currently using or have used HIV pre-exposure prophylaxis (PrEP) as part of their HIV prevention strategy for less than 1 month.

**Figure 1:**
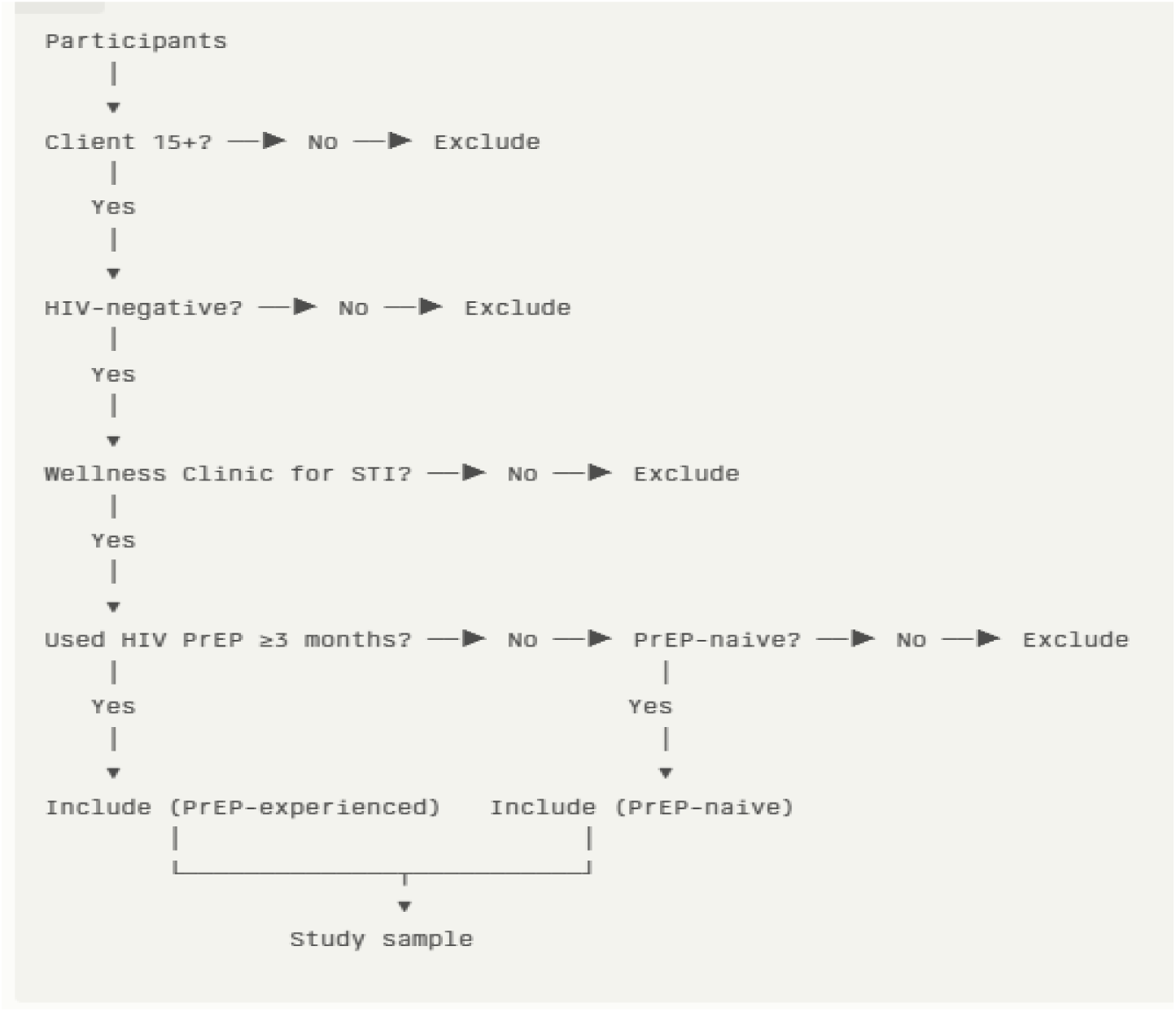
Participants’ recruitment flow chart.

### 4.7. Laboratory testing

Clients attending the wellness clinic will undergo a series of tests. Blood, urine, and genital swab samples will be collected from the clients. HIV status will be determined using rapid HIV tests (HIV self-testing kit, Determine or Unigold). Syphilis will be detected using the Treponemal rapid test or the RPR (Rapid Plasma Reagin) test. Hepatitis B will be tested with rapid antigen tests. Gonorrhea and chlamydia will be tested using GeneXpert nucleic acid amplification tests (NAATs) (if available) on urine samples (for males) and genital swabs (for females). Rapid STI tests will be used as an alternative to detect Gonorrhea, Chlamydia and Trichomonas vaginalis. If available. All tests will follow the manufacturer’s guidance and the Standard Operating Procedures. These tests are routinely performed as part of the Wellness Clinic.

### 4.8. Data management

#### 4.8.1. Data source, mining, and extraction

Data for the study will be obtained from an interviewer-administered survey. The survey questions are developed based on the study objectives and guidance from the World Health Organisation on the conduct of STI surveys (**Supplementary File 1**). The survey is divided into sections – demographic characteristics, health information, sexual behaviour, frequency and consistency of condom use, adherence to PrEP, and STI test results. The survey will be available in English and Siswati. Before the actual data collection, it will be piloted by the researchers on individuals who will not form part of the study participants. This is to ensure consistency and accuracy. The survey will be administered in a private room for confidentiality.

Patient records available in the Client Management Information System (CMIS) will be the second source of data for the study. The CMIS is the main Electronic Medical Record (EMR) widely used in Eswatini and implemented by the Ministry of Health. The STI wellness clinic registers will supplement the CMIS. The research team will check for completeness, accuracy, and relevance.

#### 4.8.2. Study Variables

The primary study variables will include demographic variables such as age, gender, sexual orientation, socioeconomic status and any other relevant demographic information; condom use patterns measures such as frequency of condom use during sexual intercourse, consistency of condom use (e.g., always, sometimes, never) and types of sexual activity and condom use in those contexts; the prevalence of specific STIs (e.g., chlamydia, gonorrhoea, trichomonas, syphilis) and the overall STI rates in the population studied; HIV PrEP Usage variables (whether participants are current users of PrEP and duration of PrEP use) and the HIV test results.

Other variables which will enrich the analysis and provide a more comprehensive understanding of the subject include frequency and type of Sexual Partners (regular or casual); risk perception and attitudes (risk of HIV and other STIs, effectiveness of PrEP and condoms, personal beliefs about STI prevention); knowledge of STI and HIV prevention; health behaviour variables (alcohol use, drug use, regular STI screening); Previous STI history (history of STI diagnoses and recurrence of STIs); educational background and level of education; Socioeconomic factors such as employment status, income level and living conditions; marital status.

#### 4.8.3. Data analysis plan

All data will be entered into an Excel spreadsheet to enable linkage between the different data sources. Data will be checked for completeness, accuracy and relevance. Data will be cleaned to remove any inconsistencies or incomplete records. Regular backups will be scheduled weekly, depending on the volume of data. Frequency tables and graphs will describe categorical variables and proportions. Continuous variables will be presented using mean and standard deviation. To find whether there is a difference between groups or variables, a comparative analysis employing the T-test or Mann-Whitney test for continuous data and Pearson χ2 or Fisher’s exact test for categorical variables will be conducted. To identify the factors that influence condom use and STI rates, bivariate logistic regression analysis will be conducted for each independent variable using Stata 17 statistical software. A p-value of <0.05 will be deemed statistically significant with a 95% confidence interval.

#### 4.8.4. Confidentiality and data safety

Data collected for this study will be anonymised after all the records have been linked before being made available to the research team. Completed questionnaires and other physical records will be kept in a lockable drawer. Only authorised people can access it. Electronic health records and data will be kept in secure servers with restricted access. Different levels of access will be assigned based on roles and responsibilities. All data will be anonymised to protect patient identities. Identifiers will be removed or masked in any dataset used for analysis or reporting. Backup copies will be stored in geographically separate secure locations to protect against data loss. All research team members will be trained in human subjects research, and confidentiality agreements will be required for all personnel handling the data. Data confidentiality and safety involve:

**a. Data Storage:** de-identified electronic data will be stored on a password-protected AHF computer at AHF Eswatini. Hard copies will be stored in a locked cabinet in the PI’s office.
**b. Authorised Personnel:** Access is limited to the PI, the Co-Investigator, and designated Study Personnel who would have signed a Confidentiality and Non-disclosure Statement.
**c. Data Retention and Disposal:** Electronic data will be retained for five years post-publication, then permanently deleted. The PI will shred hard copies.

### 4.9. Ethical considerations

#### 4.9.1. Ethical Review

Ethical clearance was obtained from the Eswatini Health and Human Research Review Board (EHHRRB), under the Protocol reference number EHHRRB 206/2025.

#### 4.9.2. Consent and participation

The participant’s right to autonomy will be respected by ensuring that participation in the study is voluntary. Potential participants will be given written informed consent (translated into Siswati for those who cannot read English) to sign voluntarily, without coercion, after the study has been explained to them and all their questions answered. Participants will be given a copy of the study information sheet if they want one. Participants will be informed that they can withdraw their consent at any time during the study without any negative impact on the health services they receive. An assent form will be given to minors to obtain the child’s agreement to participate. It will be written in a way that the minor participant can understand what the research involves and their right to withdraw. Due to the sensitivity and privacy of the topic, a waiver of informed consent will be requested for minor participants.

#### 4.9.3. Potential risk to participants

Participants face several potential risks. Emotional and psychological discomfort may arise from discussing sensitive topics like STI status or HIV testing results, with possible stigmatisation related to STI and HIV diagnoses. Privacy concerns include fears about the confidentiality of personal information and the clinic’s nature and location. There is also a risk of data misuse. Additional concerns include minimal physical risks from Laboratory testing procedures, social and relational impacts from disclosure (whether voluntary or accidental), challenges with understanding informed consent due to illiteracy, and potential coercion due to seeking health services.

Mitigation strategies will involve counselling and support to address emotional distress and secure data practices to ensure confidentiality. The psychosocial support services are provided by dedicated and experienced counsellors routinely available at the AHF LaMvelase. They are offered free of charge to the participant as part of the study’s risk mitigation strategy. Transparent reporting and ethical reviews will prevent data misuse. Trained professionals will conduct STI testing, and participants will be informed about potential risks. Maintaining privacy and providing relationship support will help manage social risks. Clear communication and thorough explanations will aid informed consent, and emphasising the voluntary nature of participation will reduce coercion. These measures will ensure the ethical conduct of the research and the safeguarding of participants’ rights and well-being.

#### 4.9.4. Potential benefits to participants

Participants will benefit from all the services provided free of charge. Participants will gain valuable information about STI prevention, PrEP, and safe sexual practices, which can empower them to make informed decisions about their sexual health. They may also benefit from increased access to healthcare services, such as STI testing and counselling, leading to earlier detection and treatment of STIs and improved health outcomes. The research assistant will further explain that the study findings will add to the body of knowledge and contribute to improved service delivery by providing evidence-based care to the public.

No financial incentives will be provided to participants for this study. Participation is entirely voluntary, and participants are compensated only through the standard of care services offered at the clinic (free STI screening and treatment, PrEP provision). This ensures that participation is non-coercive.

## 5. Discussion

### 5.1. Dissemination and utilisation of study findings

Through policy briefs, reports, meetings, and presentations, this study’s findings will be shared with key stakeholders, including national and international audiences. Abstracts will also be presented at national and international conferences to maximise the study’s impact and share evidence-based knowledge.

| **Audience** | **Method** | **Purpose** | **Timing** |
| --- | --- | --- | --- |
| **Ministry of Health (MOH)** | -Written study report<br>-Policy Briefs and Meetings | To inform policy updates and recommendations. | 2-3 months post-study |
| **AHF Africa Bureau and the global AHF Bureau** | - Reports<br>- Strategic Meetings<br>-PowerPoint presentation | - Discuss findings and implications.<br>-Update on impact and align global strategies.<br>Advocacy | 2-3 months post-study |
| **AHF Staff** | Internal Workshops/CMEs | Share findings and improve internal practices. | 1-2 months post-study |
| **HIV Care & Treatment TWG, Support partners** | -PowerPoint presentation<br>- Training, Sessions, Webinars,<br>- Collaborative Meetings | Share significant findings, review findings for integration into treatment guidelines | 3-4 months post-study |
| **Healthcare workers** | -PowerPoint presentation | Share significant findings and enhance | 2-4 months post-study |
|  | -ReHSAR, NaHSAR, National conferences, Training Sessions, Webinars | understanding and application of findings. |  |

#### Results Utilisation Plan

Findings will be used to:

1. Inform Targeted Counselling at the clinic;
2. Guide MoH Policy on national PrEP programming and STI screening; and
3. Improve Service Delivery by refining resource allocation.

### 5.2. Strengths, Limitations and Mitigation Plan

#### 5.2.1. Strengths

The study addresses a significant gap in research by investigating the relationship between PrEP use and condom use patterns in Eswatini. Given that few studies have explored this dynamic in the country, this research is crucial for understanding how PrEP impacts sexual health behaviours and STI incidence, highlighting the importance of context-specific data to refine and optimise prevention strategies. This analysis is also crucial, given the mixed findings from other studies regarding PrEP’s impact on condom use. The study setting is a high-volume facility in the central part of the Manzini region and one of the two most densely populated administrative regions in Eswatini. Therefore, this could enhance the generalizability of the study findings. Moreover, the use of a specialised STI clinic as the study setting enhances the reliability of the data on STI prevalence and sexual health behaviours.

#### 5.2.2. Limitations

The potential for sexual risk compensation, where individuals might reduce condom use due to a perceived decrease in HIV risk (Freeborn & Portillo, 2018), could complicate the interpretation of results. Selection bias may be introduced due to the use of purposive sampling, but also because Individuals seeking care at such a clinic could have different behaviours and risk profiles compared to those in broader or less specialised healthcare settings, potentially impacting the generalizability of the findings. The reliance on self-reported data for condom use and sexual behaviour might introduce recall and social desirability biases. Moreover, this study will use a cross-sectional design; therefore, causality cannot be established since data will be collected at one point in time. This study will be facility-based; hence, the results of this study may not be generalisable to other settings or populations.

## 6. Timelines

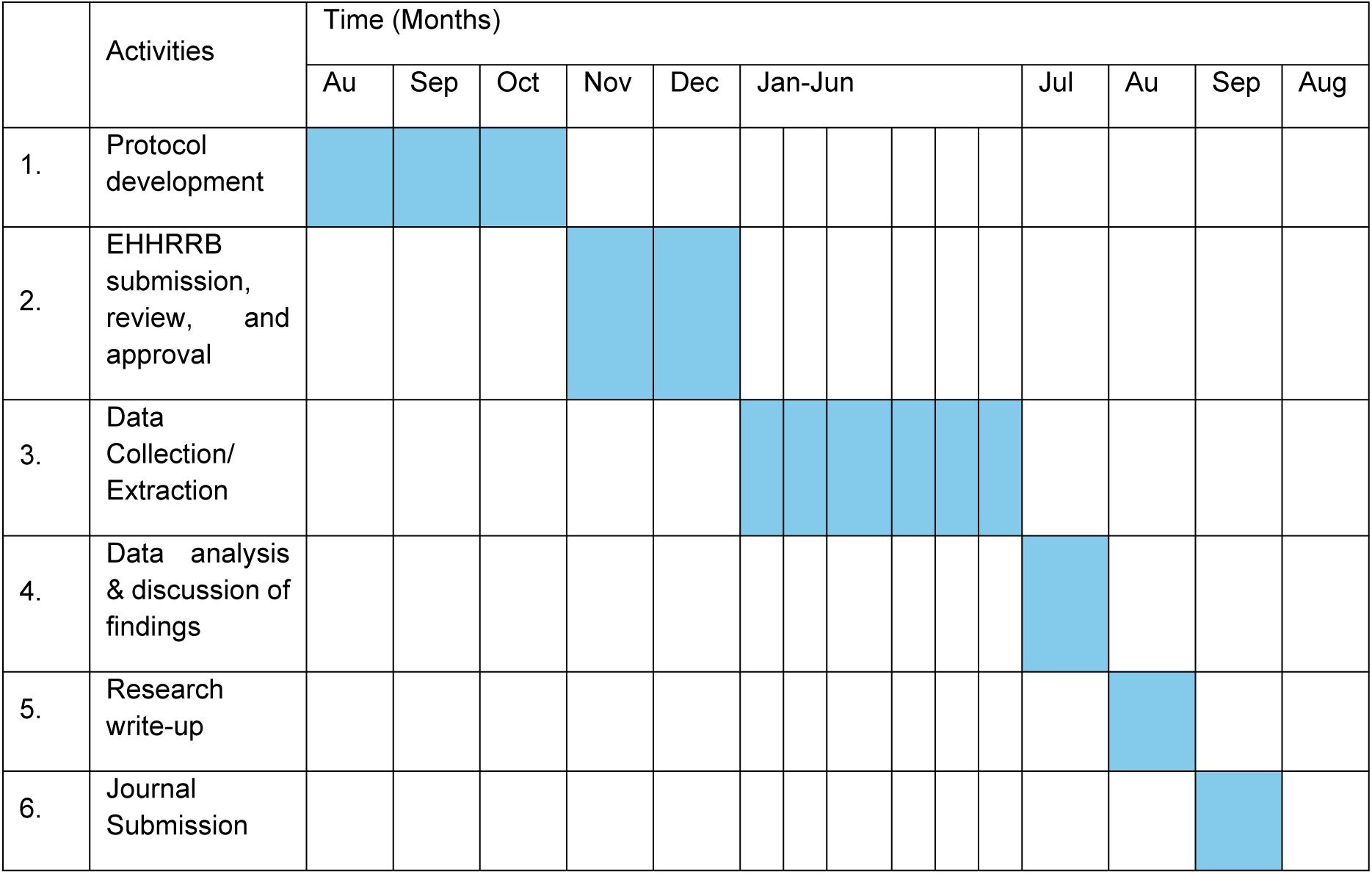

## 7. Authors’ Contributions

YM conceptualised the study, developed the research protocol, and oversees implementation and ethical compliance. PN and KLM contributed to protocol writing, including review of study tools and consent materials. VW and SN reviewed the protocol, methodology, ensured compliance with research standards, and leads data management and analysis. NN provided input on study design and protocol refinement. SG, NM, BD, NMh, and ND contributed to protocol review, translation, and implementation support.

## 8.#Acknowledgements

The authors acknowledge the contributions of the AHF Eswatini team, the LaMvelase STI Wellness Clinic staff, and all study personnel involved in protocol development and implementation.

## 9. Supporting Information

- **Supplementary file 1:** Data Collection Tool (English and Siswati)
- **Supplementary file 2:** Participant Information Sheet
- **Supplementary file 3:** Consent Form
- **Supplementary file 4:** Confidentiality and Non-disclosure Statement
- **Supplementary file 5:** Conflict of Interest Statement

## Metadata

- **Study type:** Prospective cross-sectional quantitative study
- **Setting:** LaMvelase STI Wellness Centre, Manzini, Eswatini
- **Study period:** February-June 2026
- **Population:** HIV-negative individuals aged ≥15 years attending STI services
- **Ethics approval:** EHHRRB 206/2025
- **Registration:** Not applicable (observational study)

## Data Availability

All the relevant data that will be generated or analysed during this study will be available upon request, and subject to ethical approval and confidentiality requirements. De-identified data may be shared for research and public health purposes in accordance with regulatory standards.

## Acronyms

AHF: AIDS Healthcare Foundation
ART: Antiretroviral Therapy
CDC: Centres for Disease Control and Prevention
CMIS: Client Management Information System
CT: Chlamydia trachomatis
EHHRRB: Eswatini Health and Human Research Review Board
EMR: Electronic Medical Record
FDA: Food and Drug Administration
FTC: Emtricitabine
HIV: Human Immunodeficiency Virus
MOH: Ministry of Health
NAATs: Nucleic Acid Amplification Tests
NG: Neisseria gonorrhoea
NaHSAR: National HIV Semi-Annual Review
PEP: Post-Exposure Prophylaxis
PI: Principal Investigator
POCT: Point of Care Testing
PrEP: Pre-exposure Prophylaxis
ReHSAR: Regional HIV Semi-Annual Review
RPR: Rapid Plasma Reagin
SHIMS: Swaziland HIV Incidence Measurement Survey
SPSS: Statistical Package for the Social Sciences
STI: Sexually Transmitted Infection
STATA: Statistical Analysis Software
TDF: Tenofovir Disoproxil Fumarate
TV: Trichomonas Vaginalis
VMMC: Voluntary Medical Male Circumcision
WHO: World Health Organization
3TC: Lamivudine

